# AI-based synthetic simulation CT generation from diagnostic CT for simulation-free workflow of spinal palliative radiotherapy

**DOI:** 10.1101/2025.09.02.25334595

**Authors:** Yiding Han, Alexander Nicola Hanania, Zaid Ali Siddiqui, Vincent Ugarte, Boran Zhou, Abdallah S.R. Mohamed, Piyush Pathak, Daniel Allen Hamstra, Baozhou Sun

## Abstract

**Purpose/Objective:** Current radiotherapy (RT) planning workflows rely on pre-treatment simulation CT (sCT), which can significantly delay treatment initiation, particularly in resource-constrained settings. While diagnostic CT (dCT) offers a potential alternative for expedited planning, inherent geometric discrepancies from sCT in patient positioning and table curvature limit its direct use for accurate RT planning. This study presents a novel AI-based method designed to overcome these limitations by generating synthetic simulation CT (ssCT) directly from standard dCT for spinal palliative RT, aiming to eliminate the need for sCT and accelerate the treatment workflow.

**Materials/Methods:** ssCTs were generated using two neural network models to adjust spine position and correct table curvature. The neural networks use a three-layer structure (ReLU activation), optimized by Adam with MSE loss and MAE metrics. The models were trained on paired dCT and sCT images from 30 patients undergoing palliative spine radiotherapy from a safety-net hospital, **with** 22 cases used for training and 8 for testing. To explore institutional dependence, the models were also tested on 7 patients from an academic medical center (AMC). To evaluate ssCT accuracy, both ssCT and dCT were aligned with sCT using the same frame of reference rigid registration on bone windows. Dosimetric differences were assessed by comparing dCT vs. sCT and ssCT vs. sCT, quantifying deviations in dose-volume histogram (DVH) metrics, including Dmean, Dmax, D95, D99, V100, V107, and root-mean-square (RMS) differences. The imaging and plan quality was assessed by four radiation oncologists using a Likert score. The Wilcoxon signed-rank test was used to determine whether there is a significant difference between the two methods.

**Results:** For the safety-net hospital cases, the generated ssCT demonstrated significantly improved geometric and dosimetric accuracy compared to dCT. ssCT reduced the mean difference in key dosimetric parameters (e.g., Dmean difference decreased from 2.0% for dCT vs. sCT to 0.57% for ssCT vs. sCT with significant improvement under the Wilcoxon signed-rank test) and achieved a significant reduction in the RMS difference of DVH curves (from 6.4% to 2.2%). Furthermore, physician evaluations showed that ssCT was consistently rated as significantly superior for treatment planning images (mean scores improving from “Acceptable” for dCT to “Good to Perfect” for ssCT), reflecting improved confidence in target and tissue positioning. In the academic medical-center cohort—where technologists already apply meticulous pre-scan alignment—ssCT still yielded statistically significant, though smaller, improvements in several dosimetric endpoints and in observer ratings.

**Conclusion:** Our AI-driven approach successfully generates ssCT from dCT that achieves geometric and dosimetric accuracy comparable to sCT for spinal palliative RT planning. By specifically addressing critical discrepancies like spine position and table curvature, this method offers a robust approach to bypass the need for dedicated sCT simulations. This advancement has the potential to significantly streamline the RT workflow, reduce treatment uncertainties, and accelerate time to treatment, offering a highly promising solution for improving access to timely and accurate radiotherapy, especially in limited-resource environments.

## Introduction

Over half of cancer patients undergo radiotherapy (RT) during their treatment course, and this demand continues to grow due to the increasing cancer incidence in an aging population, as well as broader applications of RT [1]. The advancement and adoption of sophisticated techniques such as stereotactic radiotherapy, image-guided radiotherapy, and the integration of radiotherapy with immunotherapy have significantly increased both the utilization and complexity of modern RT practices. Delays in RT initiation are strongly associated with poorer clinical outcomes, including decreased local control and increased disease progression, underscoring the critical need to streamline the treatment pathway [2]. Palliative RT, which represents a significant portion of all radiotherapy indications, is crucial for alleviating symptoms, improving quality of life, and occasionally extending survival in patients with bone and soft-tissue metastases [3]. Spinal metastases, in particular, often cause functional deficits due to severe back pain and, in some cases, spinal cord compression [4]. Timely intervention of spinal RT is essential for pain relief and neurological symptom management, directly impacting patients’ quality of life.

Successful RT delivery necessitates meticulous coordination among multidisciplinary team members through clearly defined and carefully managed processes, from initial consultation and treatment planning to the precise administration of radiation doses, highlighting the importance of streamlined clinical workflows. A critical and often time-intensive step is the acquisition of a dedicated simulation CT (sCT). sCT serves as the anatomical reference for treatment planning and patient setup during delivery. However, sCTs have less wide availability compared with diagnostic CT (dCT), and requires a separate, in-person patient visit using specialized equipment and positioning (e.g., flat treatment couch geometry, specific immobilization) to accurately replicate the treatment position [5]. This necessity for a dedicated sCT scan contributes significantly to treatment initiation delays, consuming valuable time and resources for both patients and healthcare providers [6-12].

To mitigate sCT-related delays, alternative approaches such as Diagnostic Scan-Based Planning (DSBP) have been explored, leveraging existing dCT images [13-17]. Prospective studies have shown DSBP substantially reduced wait time for treatment initiation without detriment in plan deliverability or quality [18, 19]. These studies do not compare dCT-based delivery with sCT delivery to robustly measure dosimetric differences. Similarly, while synthetic CT generation from other modalities like MRI and CBCT has shown promise in some areas [20, 21], the specific challenge of generating synthetic *simulation* CT from *diagnostic* CT that accurately replicates sCT geometry, particularly addressing differences in patient positioning and table curvature, has received less attention, especially using AI-based transformation approaches.

In this study, we propose to evaluate **a novel AI-based framework designed to generate synthetic simulation CT (ssCT) from standard dCT scans for spinal palliative RT**. Our approach employs machine learning to correct for spine position misalignment and table curvature differences, aiming to produce ssCT images suitable for accurate treatment planning without requiring a separate sCT scan. We developed this AI-based framework by converting dCT to ssCT to improve the accuracy of dose calculation in the DSBP method. We also aim to evaluate the accuracy and clinical suitability of the generated ssCT by comparing its geometric correspondence and dosimetric outcomes against standard dCT with rigid registration to sCT, which serves as our ground truth reference. Finally, we aim to assess ssCT performance by qualitiative physician acceptance evaluation criteria. The model was trained and validated on data from a safety-net hospital, then independently tested on an external cohort from an academic medical center (AMC).

## Method and materials

### A. AI framework

The AI framework developed for generating ssCT from dCT comprises two main components to correct geometric discrepancies inherent between dCT and sCT acquisitions: (1) spine position adjustment and (2) table curvature adjustment. Despite rigid registration based on bone windows, which simulates couch adjustments during treatment, the spine often remains misaligned due to the absence of immobilization devices commonly used during sCT scanning. Our spine position algorithm predicts sCT spine positioning from dCT data, thereby preventing mis-contouring of the spine and PTV in DSBP. Additionally, as dCTs are typically acquired on a curved tabletop while sCTs use a flat tabletop, the table curvature adjustment algorithm corrects the body’s transverse lower contour, aligning it with sCT setup. These adjustments minimize contour discrepancies and dose calculation discrepancies arising from differences in the external body contour between dCT and sCT.

#### A.1. Spine position adjustment

The spine position adjustment algorithm begins by identifying a common reference point, the heart center point. on the dCT. Two vectors are defined: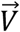, extending from the heart center point to the spinal cord center of the dCT, and 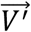, extending from the heart center point to the spinal cord center of the sCT. We then calculate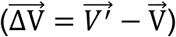, the 3D displacement to map the dCT spinal cord position to the sCT spinal cord position, as shown in Figure 1. A neural network is employed, with the heart center point, 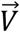 and z position as the input, and 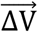 as the output. The neural network structure is: first with 64 neurons and the second with 128 neurons, both using the ReLU activation function. A third hidden layer with 64 neurons, also with ReLU activation. The model is compiled using the Adam optimizer, with mean squared error (MSE) as the loss function to minimize the difference between predicted and actual shifts, and mean absolute error (MAE) as an additional metric to evaluate prediction accuracy.

**Figure 1.**
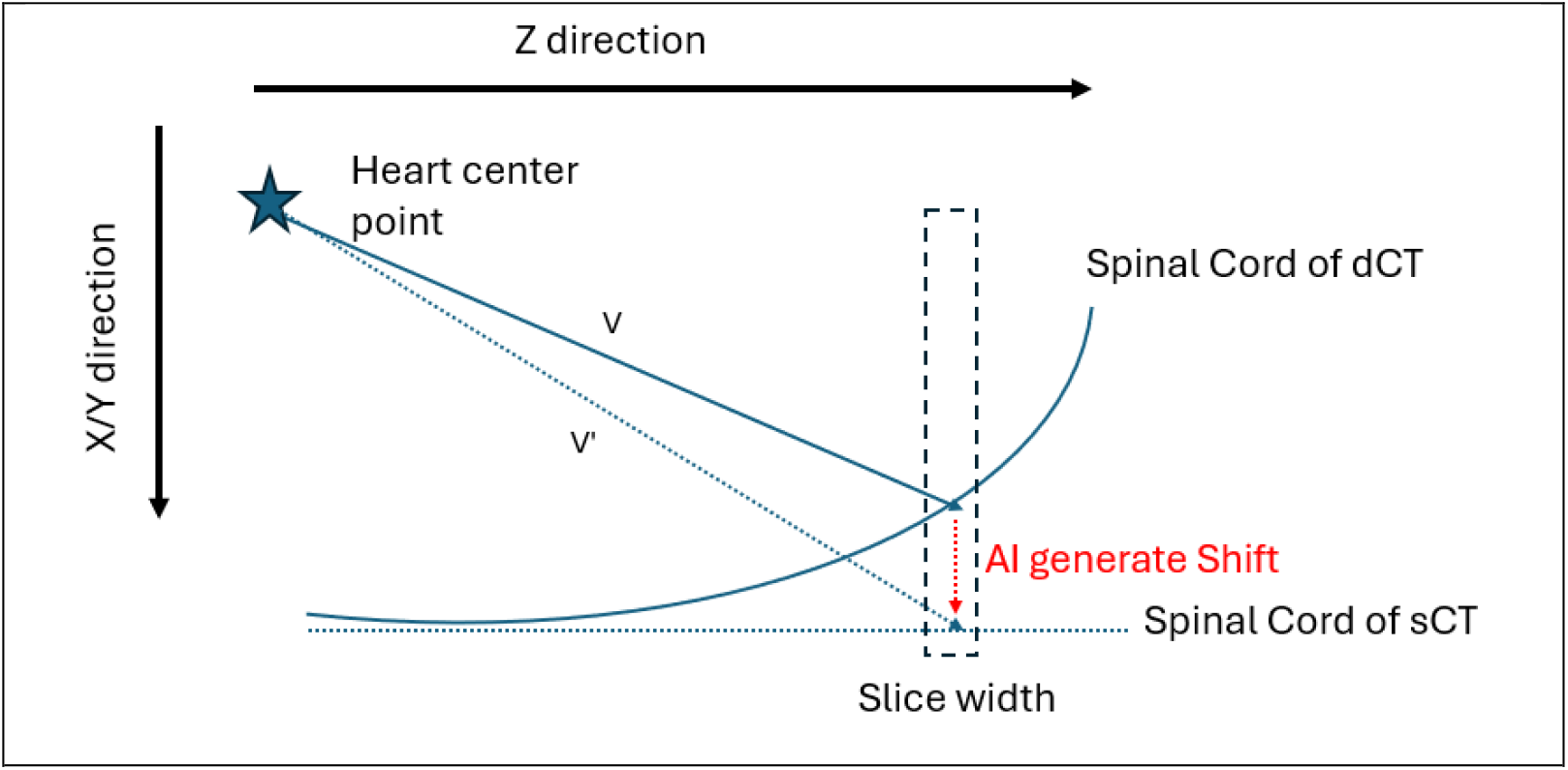
Spine position Adjustment

### A.2. Table curvature adjustment

The AI framework’s table curvature adjustment corrects the body’s lower contour to align dCT with sCT setup. The process begins by using the center of the spinal cord in each slice along the Z-axis as the reference point, following the spine position adjustment. For each slice, points on the body external contour of dCT (after spine position adjustment) and sCT are identified. A set of angles ranging from 0 to 180 degrees is used to interpolate points on dCT and sCT boundaries, with the spinal cord center as the origin. The interpolation ensures a smooth transition between the curved dCT contour and the flatter sCT contour, as illustrated in Figure 2. A neural network facilitates this adjustment, taking as inputs the points on dCT body external contour, the slice Z position, and the heart center point, and producing the corresponding points on sCT body external contour as outputs. The neural network configurations for the table curvature adjustment are the same as the spine position adjustment, except for the input and output.

**Figure 2.**
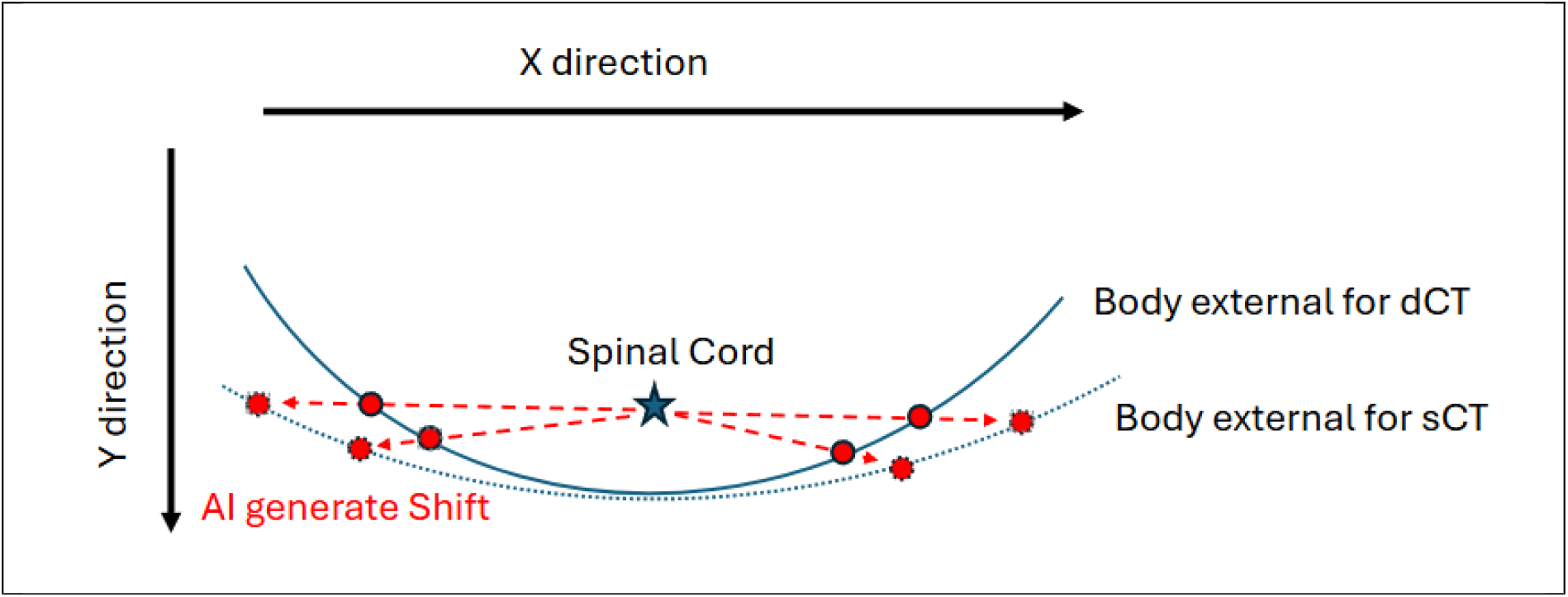
Table Curvature Adjustment

### Data preparation

This study involved 30 consecutive spinal palliative RT patients at a safety-net institution. Of these, 60% received volumetric modulated arc therapy (VMAT), and 40% received 3D conformal radiotherapy (3D RT). The AI framework was trained and internally validated using 22 patients, with 8 hold-out patients reserved for accuracy evaluation. To examine potential institution-specific effects, 7 additional cases from an AMC in the same health-care network were included. Patient selection was based on two criteria: (1) diagnostic imaging fully encompassing the planning target volume (PTV) as defined during simulation, and (2) consistent body shape between diagnostic and simulation imaging, as significant weight loss between procedures could alter dose calculations due to changes in fat and muscle composition. The typical interval between dCT and sCT was less than 2 weeks.

For each patient, a dCT and sCT with an existing treatment plan were paired and aligned using frame of reference automated rigid registration based on bone window kernel in RayStation 11B (RaySearch Laboratories, Stockholm, Sweden). sCTs included pre-existing manual segmentations performed by the treating radiation oncologist. The heart and spinal cord segmentations on dCTs were segmented automatically using MVision AI Contour+ software (MVision AI, Helsinki, Finland).

For each patient, the trained AI framework generates a ssCT from the dCT in the same frame of reference without incorporating any information from the sCT. A treatment plan, developed using the sCT with its corresponding segmentation, is then applied dCT, sCT, and ssCT to evaluate dosimetric differences across these imaging modalities. The ssCT and dCT will use the same frame of reference as mentioned before.

### Evaluation metrics

Dosimetric parameters were extracted to compare the plans, including Dmax, D95, D99, V100, and V107. The root-mean-square (RMS) difference in each parameter was calculated to assess variations in the dose-volume histogram (DVH) curves of the PTV across the plans as shown below. D_1_(*i*) and *D*_2_(*i*) are the doses at the i-th volume point for the two curves.

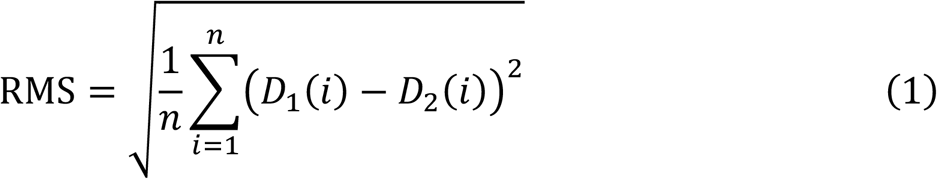

Four physicians—three experienced radiation oncologists and one fifth-year (PGY-5) radiation oncology resident—evaluated dCT and ssCT plans on a scale of 1 to 5 (5: Perfect, 4: Good, 3: Acceptable, 2: Unacceptable, 1: Poor) based on target and tissue positioning and dose distribution comparing the plans to the sCT plans as the golden standard.

### Statistical Analysis

Because every patient produced paired error values from the two pipelines (dCT and ssCT) and the error distributions were visibly non-normal, we assessed performance with the Wilcoxon signed-rank test, the non-parametric analogue of the paired t-test. For each dosimetric metric we defined the paired difference:

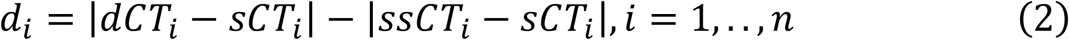

A positive *d*_*i*_ Indicates that the dCT error exceeds the ssCT error for that patient.

Paris with *d*_*i*_ = 0 (uninformative about direction) were removed, leaving n′ non-zero differences.

Their magnitudes |*d*_*i*_| were ranked (ties assigned mid-ranks); the signed-rank statistic

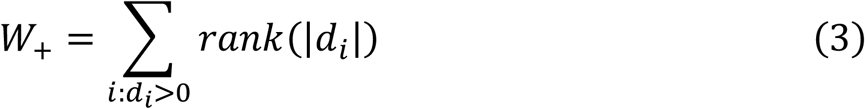

is the sum of ranks attached to positive differences.

Under the null hypothesis of no systematic difference, every non-zero pair is equally likely to be positive or negative; hence all 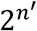 sign permutations are equiprobable.

Because of the limited number of cases (n’<20) for all metrics, the exact one-sided p-value was obtained from the exact null distribution of *W*_+_:

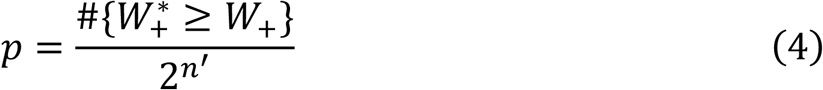

Where 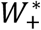 is the rank sum generated by each possible sign permutation.

The alternative hypothesis was directional—that ssCT yields smaller errors than dCT (means ssCT performs better than dCT)—so only the upper tail of the distribution was used. Statistical significance was declared for p < 0.05.

### Observer-agreement analysis

Inter-observer variability of the four radiation oncologists’ scoring was quantified in two complementary ways:

#### Simple agreement

- **Exact match (%)** – for each of the six rater–pair combinations (*C*_4_,2) the proportion of targets with identical scores was calculated and then averaged over the six pairs.
- **≤ 1-step agreement (%)** – the same procedure, but a pair was counted as agreeing when their scores differed by at most 1 point (e.g. 4 vs 5).

#### Chance-corrected agreement: Gwet’s AC^2^ (quadratic weights)

For an ordinal scale with *k = 5* categories, the quadratic weight matrix is

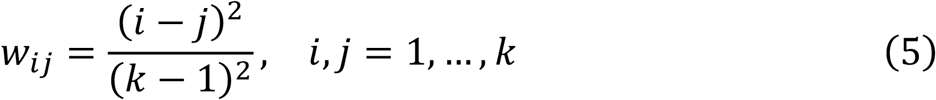

Let *o*_*ij*_ be the observed proportion of rater-pair ratings falling in cell (i,j) of the *k × k* confusion matrix and *p*_*i*_ the marginal category proportion.

The observed and expected disagreements are

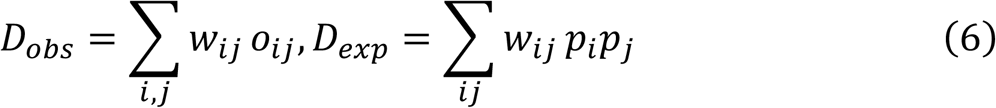

and Gwet’s coefficient is

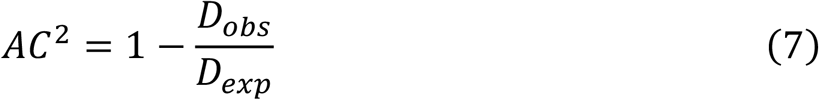

The exact sampling distribution was used (no large-sample approximation), and interpretation followed the conventional scale: < 0 = poor, 0–0.20 = slight, 0.21–0.40 = fair, 0.41–0.60 = moderate, 0.61–0.80 = substantial, > 0.80 = almost perfect agreement.

## Results

Figure 3 depicts the patient characteristics in the safety-net and AMC cohorts, with separate panels showing (upper left) the frequency of treated vertebral levels, (upper right) the distribution of primary tumor histologies, (lower left) the age distribution, and (lower right) the body-mass index (BMI) distribution.

**Figure 3.**
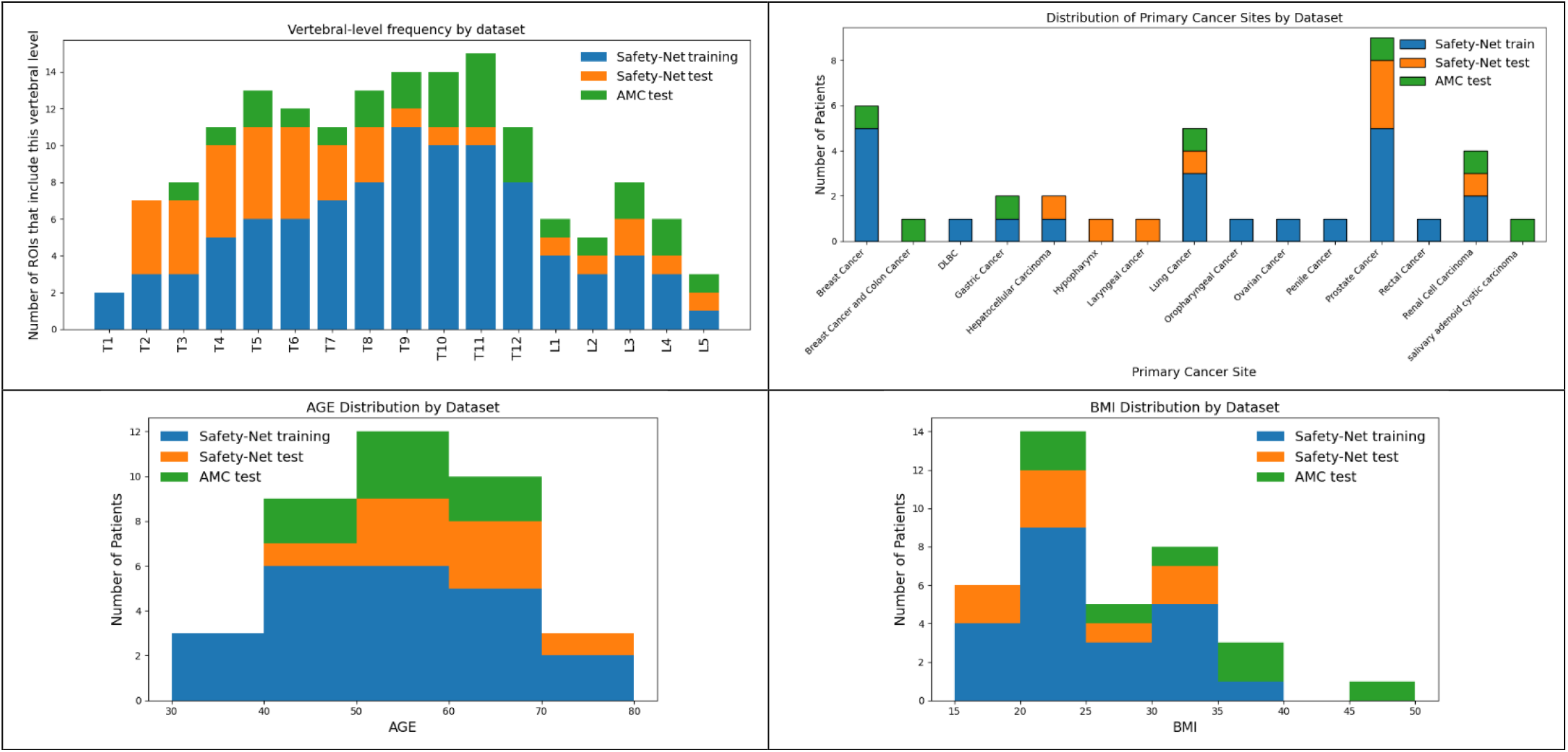
Patient demographics for the training (blue) and test (yellow) datasets from the Safety-net institution and external test dataset (green) from the AMC

**Figure 1.**
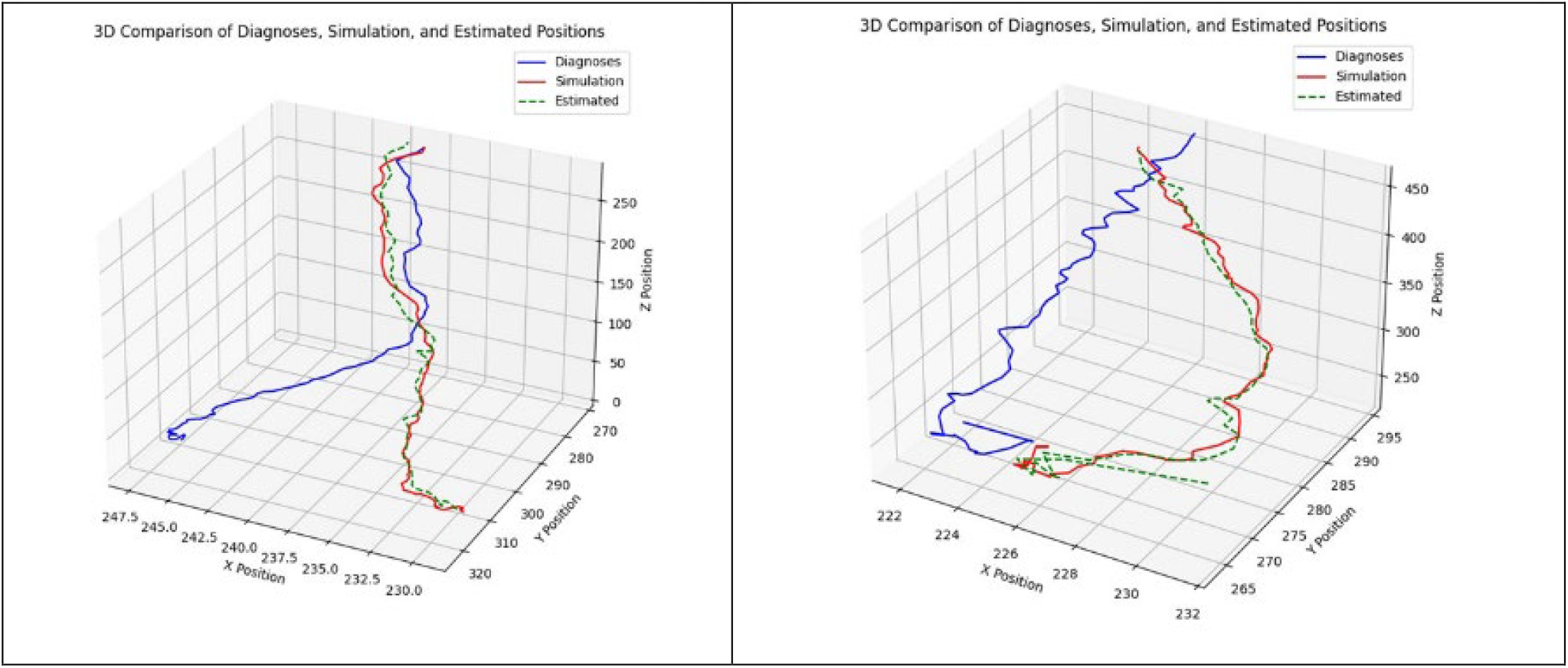
Two examples of spinal cord position adjustment results

**Figure 2.**
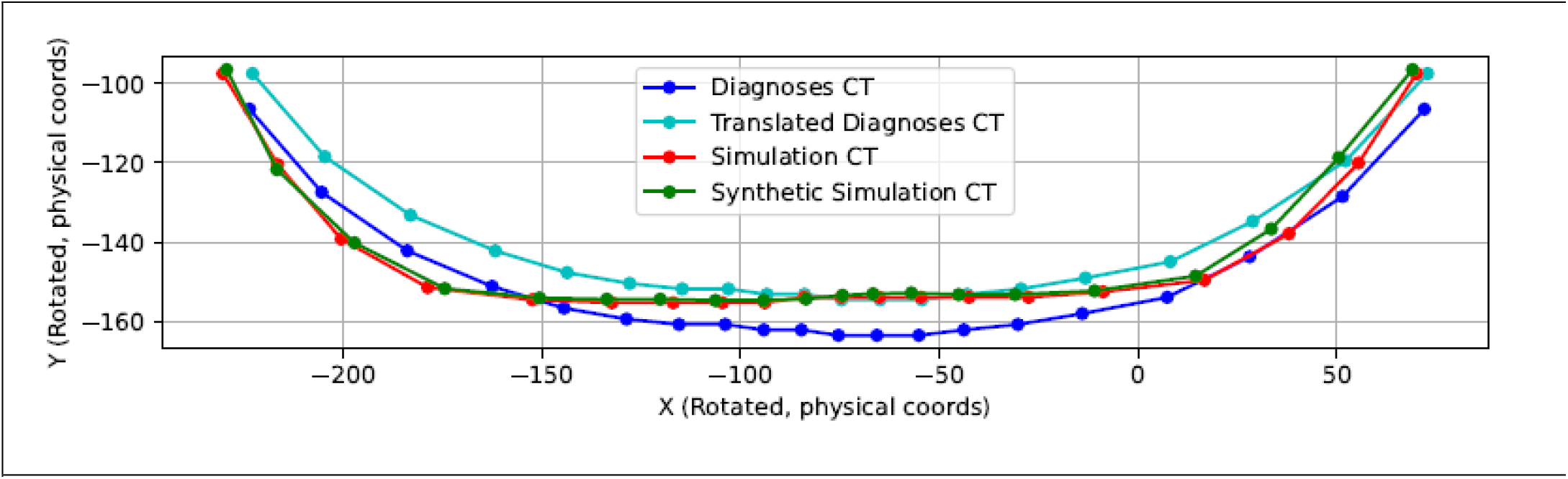
Table curvature adjustment result

**Figure 3.**
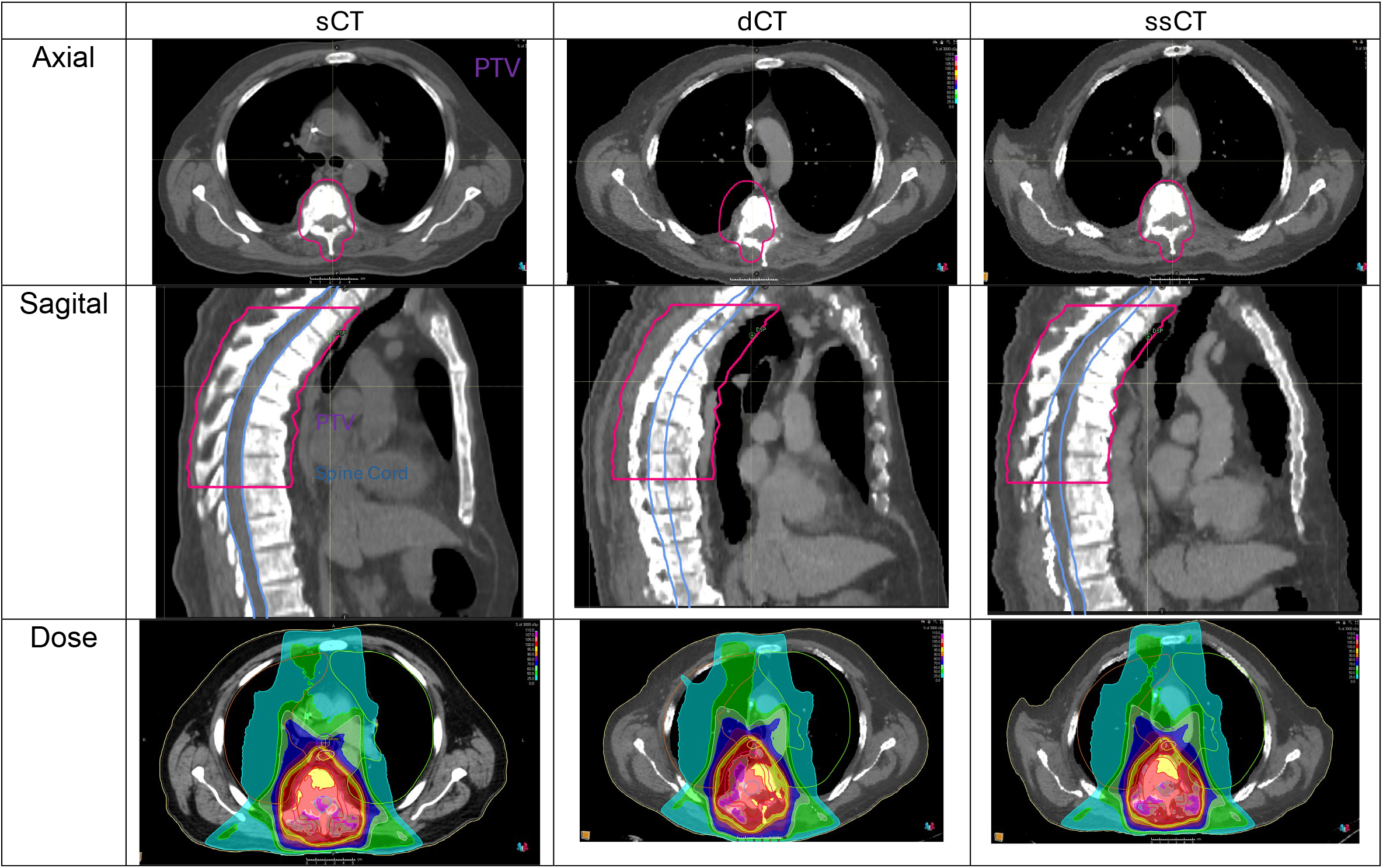
A patient example of sCT, dCT, and ssCT with PTV contouring

Figure 4 presents two examples of the spinal position adjustment results on testing dataset. The blue line depicts the center of the spinal cord in dCT across various Z positions (slices), the red line indicates the center of the spinal cord in sCT, and the green dashed line shows the adjusted spinal cord position on dCT after applying the spine position adjustment.

Figure 5 presents the results of the table curvature adjustment. The blue line represents the body external contour in dCT, the cyan line depicts dCT body external contour after spinal position adjustment, the red line indicates the body external contour in sCT, and the green dashed line shows the final dCT body external contour following both spinal position adjustment and table curvature adjustment.

Figure 6 displays a patient case comparing sCT, dCT, and ssCT from left to right columns. The first row presents the transverse plane at a consistent Z position, with the PTV contoured in pink. The second row depicts the sagittal plane at a consistent × position, showing the contours of PTV (pink) and spinal cord (blue). The PTV and spinal cord are segmented on sCT and applied to dCT and ssCT with frame of reference registration to show the spine position deviation between dCT and sCT/ssCT. The third row illustrates the dose distributions of the different CTs with the same treatment plan at the same Z position as the first row.

Table 1 presents the average differences in dosimetric parameters between dCT and ssCT compared to sCT as the reference, using the same treatment plan.

**Table 1.**
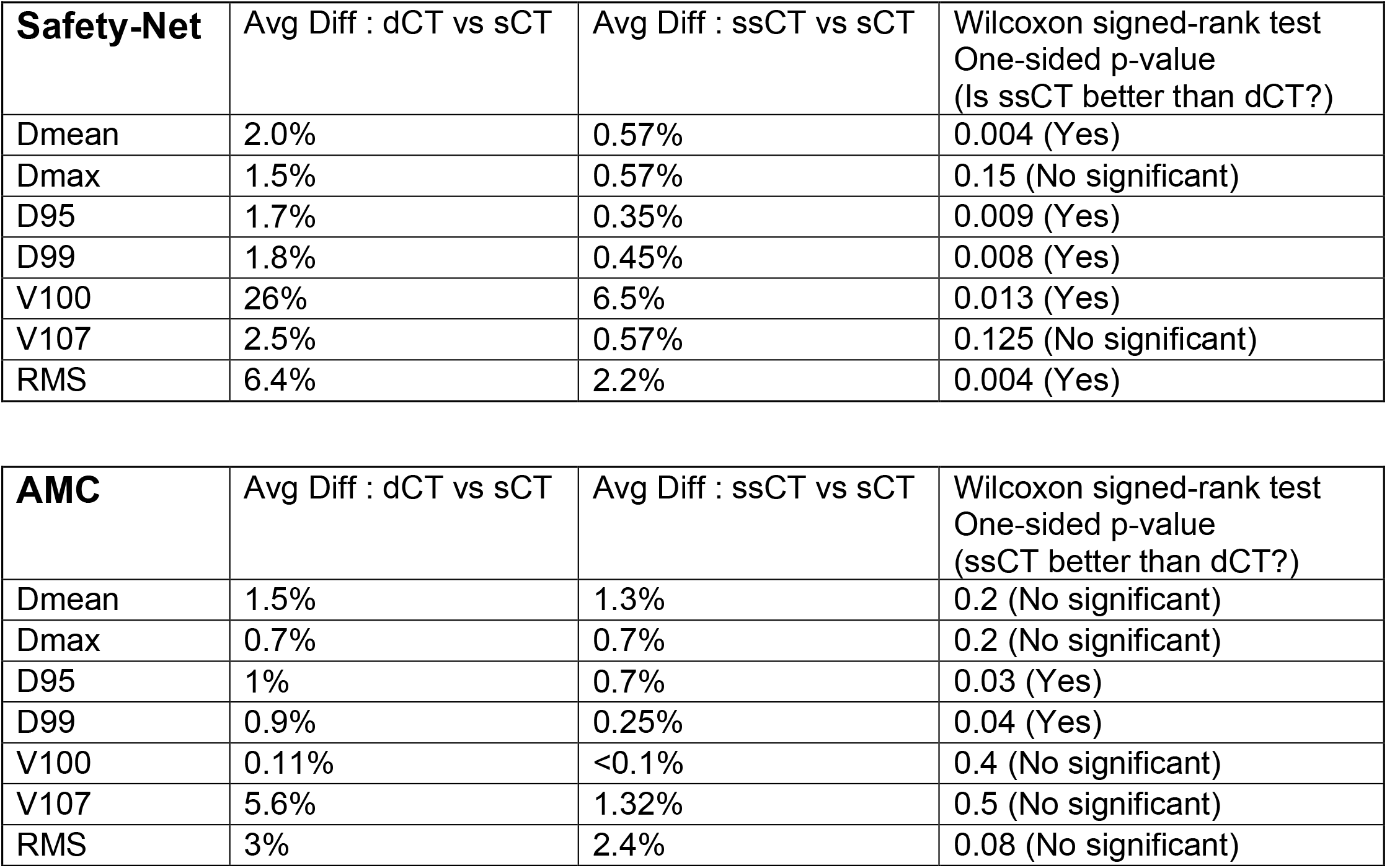
Dosimetric parameters difference of PTV for patients from two institutions.

| **Safety-Net** | Avg Diff : dCT vs sCT | Avg Diff : ssCT vs sCT | Wilcoxon signed-rank test<br>One-sided p-value<br>(Is ssCT better than dCT?) |
| --- | --- | --- | --- |
| Dmean | 2.0% | 0.57% | 0.004 (Yes) |
| Dmax | 1.5% | 0.57% | 0.15 (No significant) |
| D95 | 1.7% | 0.35% | 0.009 (Yes) |
| D99 | 1.8% | 0.45% | 0.008 (Yes) |
| V100 | 26% | 6.5% | 0.013 (Yes) |
| V107 | 2.5% | 0.57% | 0.125 (No significant) |
| RMS | 6.4% | 2.2% | 0.004 (Yes) |

| **AMC** | Avg Diff : dCT vs sCT | Avg Diff : ssCT vs sCT | Wilcoxon signed-rank test<br>One-sided p-value<br>(ssCT better than dCT?) |
| --- | --- | --- | --- |
| Dmean | 1.5% | 1.3% | 0.2 (No significant) |
| Dmax | 0.7% | 0.7% | 0.2 (No significant) |
| D95 | 1% | 0.7% | 0.03 (Yes) |
| D99 | 0.9% | 0.25% | 0.04 (Yes) |
| V100 | 0.11% | <0.1% | 0.4 (No significant) |
| V107 | 5.6% | 1.32% | 0.5 (No significant) |
| RMS | 3% | 2.4% | 0.08 (No significant) |

Table 2 presents the physician scoring of dCT and ssCT as treatment planning images, using a scale from 1 to 5 (5: Perfect, 4: Good, 3: Acceptable, 2: Unacceptable, 1: Poor). The result is shown with the standard deviation.

**Table 2.**
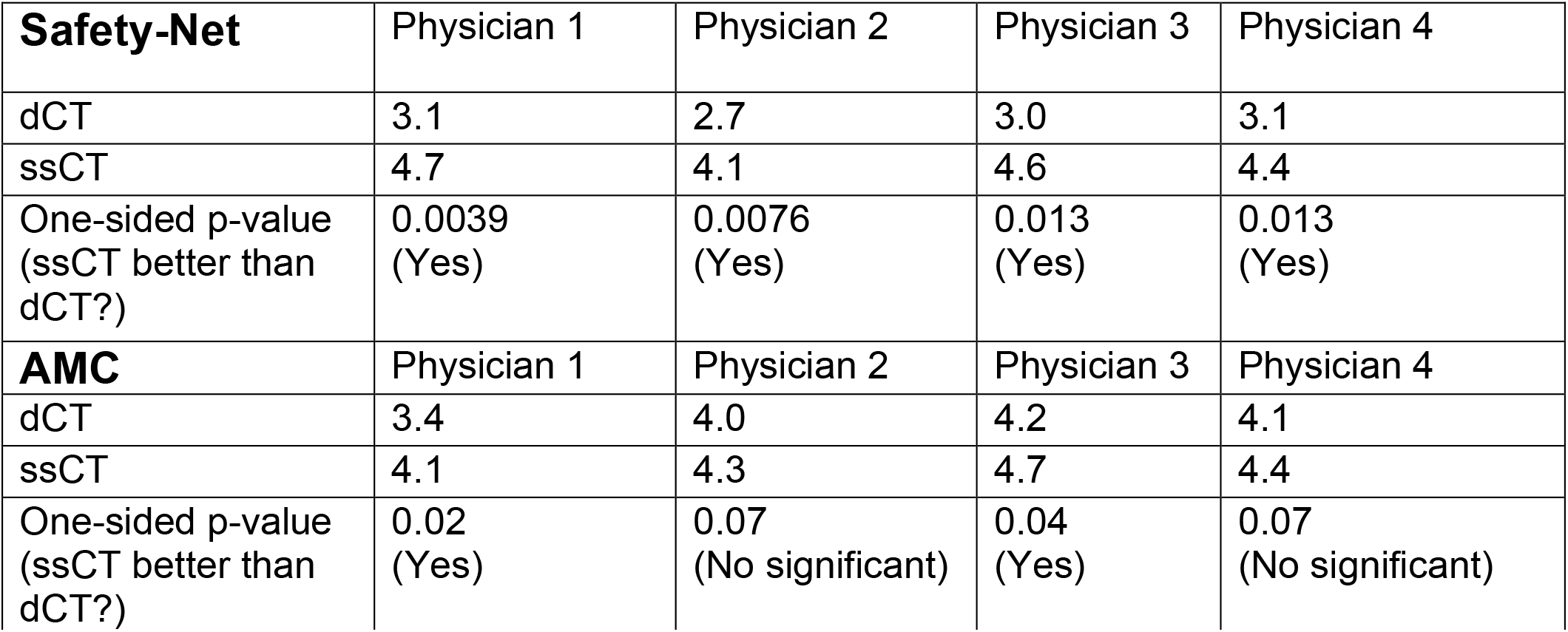
Average physician scoring for dCT and ssCT as treatment planning images.

Table 3 summarizes inter-observer agreement for the four physicians on both dCT and ssCT, based on 15 cases drawn from two institutions.

**Table 3.**
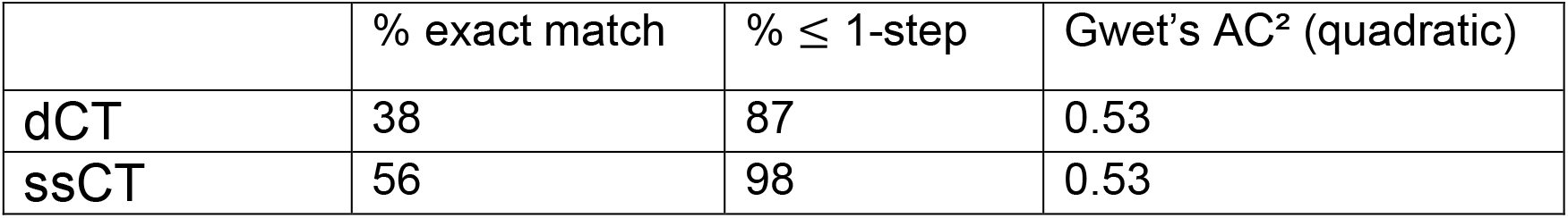
Inter-observer agreement metrics for four physicians (15 cases from two institutions).

## Discussion

Our study demonstrates that the spine position and table curvature adjustments, as demonstrated in Figures 4 and 5, show strong alignment between ssCT and sCT in terms of spinal cord positioning and body external contour, despite dCT being acquired with a different setup. The success of the model, trained on a limited dataset of 16 patients, highlights a robust correlation between dCT and sCT. This correlation is expected, as the protocols for dCT scans and RT planning are standardized within the same institution. Consequently, the model serves as a mathematical tool to capture the geometric relationship between dCT and sCT, which is inherently constrained by institutional protocols and physical factors, such as the effect of gravity on the body contour due to differing table setups (curved for dCT versus flat for sCT)

The impact of spine position and table curvature adjustments is evident in the patient case presented in Figure 6. In the transverse and sagittal planes (first and second rows), ssCT demonstrates strong alignment in spinal cord position and GTV with sCT under the treatment setup, whereas dCT exhibits a noticeable deviation in spinal cord position relative to sCT. Despite this spatial deviation, the third row reveals that the dose distribution difference between dCT and sCT is less pronounced than the spatial discrepancy. However, the dose distribution on dCT is colder on the left side of the GTV compared to sCT, indicating that using dCT for treatment planning could result in an overdosed target during delivery. This issue is mitigated in ssCT through the combined spine position and table curvature adjustments, ensuring more accurate dose delivery.

Table 1 summarizes the dosimetric comparison between the two scout pipelines, using sCT as the reference and the same treatment plan for all patients. For the safety-net hospital cohort, ssCT demonstrated a clear dosimetric advantage: mean absolute differences with respect to sCT were lower for every end-point, and the variability across patients was consistently narrower. For example, the mean-dose error (D *mean*) fell from 2.0 % for dCT to 0.57 % for ssCT (*p* = 0.004), and the global DVH deviation (RMS) declined from 6.4 % to 2.2 % (*p* = 0.004). The largest improvement was seen in target-coverage index V *100* (26 % vs 6.5 %, *p* = 0.014). These gains confirm that the spine-position and table-curvature corrections embodied in the ssCT algorithm materially enhance dosimetric fidelity in a resource-constrained environment.

In the academic medical-center (AMC) cohort the differences were smaller and, for most metrics, not statistically significant. The baseline dCT scans from the AMC already agreed well with sCT (e.g. mean DVH-RMS ≈ 3 %), reflecting the center’s stringent pre-scan alignment protocol; ssCT therefore had less room for improvement.

Qualitative image evaluation (Table 2) echoes the quantitative dosimetric results, while the inter-observer analysis in Table 3 confirms **moderate overall agreement** between physicians for both pipelines (Gwet AC^2^ = 0.53). At the safety-net institution, mean physician scores rose from *Acceptable* (2.7–3.1) for dCT to *Good–Perfect* (4.1–4.7) for ssCT, with statistically significant improvement for all four observers. At the AMC, dCT images were rated one category higher to begin with (3.4–4.2), yet ssCT still achieved a significant upgrade for two observers and maintained at least *Good* quality for the others. Thus the image-quality benefit of ssCT is most pronounced where patient positioning is sub-optimal, but remains at least equivalent under best-practice conditions.

The center-to-center disparity apparent with dCT—but not with ssCT—underscores a key operational advantage of the synthetic approach. dCT reliability depends on labour- and time-intensive alignment procedures that are difficult to sustain in high-volume, resource-limited clinics, whereas ssCT achieves comparable or superior quality by algorithmically standardizing patient geometry.

Beyond accurate spine positioning and body contour estimation, a key strength of this study is the AI model’s efficiency, which requires only 22 training cases—far fewer than conventional methods, such as 3D convolutional neural networks, which typically demand large datasets. This efficiency stems from the model’s targeted focus on spine positioning, closely correlated with the GTV, and table curvature, a primary source of deviation between dCT and sCT protocols, rather than global image details. By prioritizing local accuracy over global accuracy, the model reduces training data needs and enables computation in a CPU-only environment, eliminating the need for GPU resources. This innovation significantly enhances the method’s feasibility for resource-limited settings, broadening its potential clinical impact.

Although the spine position and couch curvature correction markedly reduced PTV misalignment, mobile soft-tissue structures—especially small, highly deformable organs such as the esophagus—remain a challenge. During blinded review, several physicians noted residual offsets between the ssCT and sCT contours for these tissues. A perfect prediction of soft-tissue location is probably unrealistic; however, generating a probabilistic envelope of likely positions would still be clinically useful for margin design.

Future investigations should focus on demonstrating that the proposed pipeline generalizes beyond the local network. Multi-institutional validation requires enrolling centers in other resource-limited settings, thereby testing the algorithm under various scanners, workflows, and patient demographics. In parallel, methodological development will shift from deterministic spine realignment to a probabilistic framework that estimates the positional envelope of mobile soft tissues. By characterizing the range rather than the exact location of organs such as the esophagus, the method could be extended to disease sites where internal target volume and gross tumor volume definition is dominated by organ motion, including pelvic, hepatic, and thoracic malignancies. These two lines of work—external validation and soft-tissue range modelling—will be essential steps toward a genuinely simulation-free planning paradigm that is both broadly applicable and resilient to variations in clinical practice.

## Conclusion

This study demonstrates the efficacy of an AI framework for generating ssCT from dCT through spine position and table curvature adjustments, addressing key challenges in RT planning for spinal palliative patients. ssCT achieved strong alignment with sCT in spinal cord positioning and body external contour despite the differing setups of dCT and sCT. Dosimetric analysis revealed that ssCT significantly outperforms dCT, reducing the mean difference in Dmean and the RMS difference in DVH curves with lower variability indicating greater stability across cases. Physician evaluations further confirmed ssCT’s superiority, with scores improving from an average “Unacceptable to Acceptable” for dCT to “Good to Perfect” for ssCT, reflecting enhanced suitability for treatment planning. These findings suggest that the proposed AI framework can effectively mitigate setup-related discrepancies, improving dosimetric accuracy and clinical reliability in RT planning. Future work should focus on validating the framework across multiple networks’ resource-limited settings, estimating soft tissue ranges to extend applicability to other disease sites like pelvic, liver, or lung cancers, and developing a generalized simulation-free RT planning approach to enhance its clinical utility and robustness.

## Data Availability

All data produced in the present study are available upon reasonable request to the authors

## Acknowledgement

We would like to thank Radiation Oncology Institute for financial support through a research grant.

## Reference

1. Atun, R., et al., Expanding global access to radiotherapy. Lancet Oncol, 2015. 16(10): p. 1153–86.

2. Robinson, D., et al., Waiting times for radiotherapy: variation over time and between cancer networks in southeast England. Br J Cancer, 2005. 92(7): p. 1201–8.

3. Radiotherapy in Palliative Cancer Care: Development and Implementation. 2012, Vienna: INTERNATIONAL ATOMIC ENERGY AGENCY.

4. Davis, D.D. and S.M. Kane, Palliation Radiation Therapy of the Spinal Cord, in StatPearls. 2025, StatPearls Publishing

5. Hegi, F., et al., 34 - Technical Requirements for Lung Cancer Radiotherapy, in IASLC Thoracic Oncology (Second Edition), H.I. Pass, D. Ball, and G.V. Scagliotti, Editors. 2018, Elsevier: Philadelphia. p. 318-329.e2.

6. Benk, V., et al., Effect of delay in initiating radiotherapy for patients with early stage breast cancer. Clin Oncol (R Coll Radiol), 2004. 16(1): p. 6–11.

7. Huang, J., et al., Does delay in starting treatment affect the outcomes of radiotherapy? A systematic review. J Clin Oncol, 2003. 21(3): p. 555–63.

8. Everitt, S., et al., High rates of tumor growth and disease progression detected on serial pretreatment fluorodeoxyglucose-positron emission tomography/computed tomography scans in radical radiotherapy candidates with nonsmall cell lung cancer. Cancer, 2010. 116(21): p. 5030–7.

9. Do, V., V. Gebski, and M.B. Barton, The effect of waiting for radiotherapy for grade III/IV gliomas. Radiother Oncol, 2000. 57(2): p. 131–6.

10. Buszek, S.M., et al., Optimal Timing of Radiotherapy Following Gross Total or Subtotal Resection of Glioblastoma: A Real-World Assessment using the National Cancer Database. Sci Rep, 2020. 10(1): p. 4926.

11. Jensen, A.R., H.M. Nellemann, and J. Overgaard, Tumor progression in waiting time for radiotherapy in head and neck cancer. Radiother Oncol, 2007. 84(1): p. 5–10.

12. Žumer, B., et al., Impact of delays in radiotherapy of head and neck cancer on outcome. Radiat Oncol, 2020. 15(1): p. 202.

13. Suzuki, K., et al., A web-based remote radiation treatment planning system using the remote desktop function of a computer operating system: a preliminary report. Journal of Telemedicine and Telecare, 2009. 15(8): p. 414–418.

14. Norum, J., et al., Telemedicine in radiotherapy: a study exploring remote treatment planning, supervision and economics. Journal of Telemedicine and Telecare, 2005. 11(5): p. 245–250.

15. Belard, A., et al., Improving Proton Therapy Accessibility Through Seamless Electronic Integration of Remote Treatment Planning Sites. Telemedicine and e-Health, 2011. 17(5): p. 370–375.

16. Glober, G., et al., Technical Report: Diagnostic Scan-Based Planning (DSBP), A Method to Improve the Speed and Safety of Radiation Therapy for the Treatment of Critically Ill Patients. Practical Radiation Oncology, 2020. 10(5): p. e425–e431.

17. Wong, S., et al., Diagnostic Computed Tomography Enabled Planning for Palliative Radiation Therapy: Removing the Need for a Planning Computed Tomography Scan. Practical Radiation Oncology, 2021. 11(2): p. e146–e153.

18. Schuler, T., et al., Real-World Implementation of Simulation-Free Radiation Therapy (SFRT-1000): A Propensity Score-Matched Analysis of 1000 Consecutive Palliative Courses Delivered in Routine Care. Int J Radiat Oncol Biol Phys, 2025. 121(3): p. 585–595.

19. O’Neil, M., et al., Diagnostic CT-Enabled Planning (DART): Results of a Randomized Trial in Palliative Radiation Therapy. International Journal of Radiation Oncology, Biology, Physics, 2024. 120(1): p. 69–76.

20. Bahloul, M.A., et al., Advancements in synthetic CT generation from MRI: A review of techniques, and trends in radiation therapy planning. Journal of Applied Clinical Medical Physics, 2024. 25(11): p. e14499.

21. Bornstein, M.M., K. Horner, and R. Jacobs, Use of cone beam computed tomography in implant dentistry: current concepts, indications and limitations for clinical practice and research. Periodontology 2000, 2017. 73(1): p. 51–72.

